# Role of *STING/TMEM173* mutation in systemic lupus erythematosus: from animal model to intrinsic human genetics

**DOI:** 10.1101/2022.12.02.22283012

**Authors:** Pichpisith Pierre Vejvisithsakul, Satima Wanachate, Pintip Ngamjanyaporn, Chisanu Thumarat, Thanitta Suangtamai, Asada Leelahavanichkul, Nattiya Hirankan, Trairak Pisitkun, Soren Riis Paludan, Prapaporn Pisitkun

**Author notes:** **CORRESPONDING AUTHOR:** Prapaporn Pisitkun, M.D., 270 RamaVI Road, Ratchathewi, Bangkok, 10400, Thailand.

## Abstract

**Objective:** We aim to confirm the function of *Sting/Tmem173* in pristane-induced lupus and identify the role of *STING/TMEM173* variants in SLE susceptibility.

**Methods:** Pristane-induced lupus model was introduced in the *Sting*-deficient mice (ENU-induced Goldenticket mutant mice). Autoantibody, histopathology, and immunophenotypes were analyzed after pristane injection for six months. Isolated DNA from 302 SLE patients and 173 healthy donors were tested for STING genotyping. We calculated the Odd Ratios of each STING variant and the inheritance patterns that significantly increased SLE susceptibility. Then, we analyzed the associations between STING genotypes and lupus phenotypes.

**Results:** The absence of STING signaling in the Goldenticket mutant mice reduced the autoantibody production and severity of glomerulonephritis in pristane-induced lupus. The human *STING* mutation at p.R284S (gain-of-function) significantly increased the SLE risk in autosomal dominant pattern [OR = 64.0860 (95%CI = 22.8605-179.6555), p < 0.0001], while the mutation at p.R232H (loss of function) reduced the SLE risk in autosomal recessive pattern [OR = 0.2515 (95%CI = 0.1648-0.3836), p < 0.0001]. The combination of STING variants in a specific inheritance pattern increased the higher OR than a single variant. The patient who had p.R284S with p.R232H showed milder disease activity than those who had p.R284S alone at the time of diagnosis.

**Conclusion:** The inhibition of STING rescued autoimmune phenotypes in pristane-induced lupus. Gain-of-function STING mutation increased SLE susceptibility and severity of the disease. These data suggested the critical function via STING-mediated signaling in SLE. Targeted at STING may provide a favorable outcome in SLE patients.

## INTRODUCTION

Systemic lupus erythematosus (SLE) is one of the most common systemic autoimmune diseases. The pathogenesis of SLE is complex, requiring the interaction between genetic susceptibility and the triggering factors in the environment. The prevalence and severity of SLE differ among genetic backgrounds (1, 2). The involvement of vital organs and treatment efficacy among ethnicities determine the outcome (3, 4). Type I interferon (IFN) is a critical cytokine that induces lupus disease. Interruption of type I IFN receptor ameliorates lupus-like phenotypes in NZB mice (5) and protects the development of pristane-induced lupus (6). The mice with *Tlr7* over-expression showed a rise in interferon-inducible genes and developed lupus phenotypes (7, 8). The nucleic sensing signaling pathways strongly initiate type I interferon production (9).

TREX1 (Three Prime Repair Exonuclease 1) degrades excess intracellular single-strand DNA to prevent the activation of interferon-stimulated genes (10). *Trex1*-deficient mice develop autoimmunity mediated through the type I IFN signaling (11). The mutations in the human *TREX1* gene cause Aicardi-Goutières syndrome and chilblain lupus (12, 13). The absence of cGAS (Cyclic GMP–AMP synthase) and Sting (Stimulator of interferon genes) signaling abrogates the lethal autoimmune phenotypes in *Trex1*-deficient mice (14, 15).

Pristane-induced lupus mice show interferon signatures and similar phenotypes with human SLE (4). IFNAR-deficient mice do not develop autoantibodies and lupus nephritis in the pristane-induced model (6). *Tlr9*-deficient C57BL/6 mice develop less histological renal injury and immune complex deposition than wild-type mice in pristane-induced lupus (16). However, *Tlr9*-deficient BALB/C mice show worsening renal pathology compared to wild-type (17). These data suggested thcriticalnt role of the mouse background in lupus development.

STING mutations are identified in patients with type I interferonopathies, so-called STING-associated vasculopathy with onset in infancy (SAVI) (18-20). The *STING* gain-of-function mutations have been identified in inflammatory lupus liked disease and familial chilblain lupus (21, 22). However, the analysis of three *STING* mutations in exonfive5 did not show an association with systemic autoimmune disease (23). The absence of STING aggravates inflammation and increases autoantibody production in the autoimmune MRL.*Fas*^*lpr*^ mice (24). Activation of STING signaling induces B cell death, and the absence of STING accelerates autoimmune arthritis in the collagen-induced arthritis model (25). Nevertheless, a previous study shows that STING signaling in dendritic cells promotes plasmacytoid dendritic cell differentiation and initiates lupus phenotypes in the 129/B6.*Fcgr2b*-deficient mice (26).

Thus, we aim to conduct a proof-of-concept study to identify whether STING is a potential targeted molecule for SLE therapy. We deployed pristane-induced lupus in the *Sting*-deficient Goldenticket mouse on a C57BL/6 background (27). The mutant mouse shows the loss of STING and type I IFN signaling function, which behaved as functional Sting-deficient mice. Pristane-induced lupus in the Sting-deficient (goldenticket or *Sting*^*gt/gt*^) mice did not develop anti-dsDNA and lupus nephritis. Also, the *Sting*^*gt/gt*^ mice limited the expansion of plasmacytoid dendritic cells after pristane injection, a similar finding to the 129/B6.Fcgr2b-deficient lupus mice (26). The *Sting*^*gt/gt*^ mice derived from the N-ethyl-N-nitrosourea-induced mutation may influence differences in autoimmune phenotypes between models. Thus, the STING inhibitor was designed to specifically target STING signaling to determine STING’s role in lupus (28). The STING antagonist (ISD017) interfering with STIM1 inhibits lupus pathology in the lupus-prone 129/B6.*Fcgr2b*-deficient mice. ISD017 also reduces the CXCL10 production from SLE patients’ peripheral blood mononuclear cells (PBMC) (28).

Single nucleotide polymorphisms (SNPs) of *STING/Tmem173* in human are common in the population and affects the immune response against viral infection (29). Here, we identified that the *STING* gain-of-function mutation increased SLE risk in humans, while *STING* loss-of-function reduced the risk. The combination of *STING* variants showed the additive risk of SLE. In summary, we confirmed that loss-of-STING function rescued autoimmune phenotypes in pristane-induced lupus and identified *STING* variants and mutations associated with the SLE risk in humans. Our data suggested that STING plays a vital role in SLE and is a potential target for therapeutic intervention.

## MATERIALS AND METHODS

### Pristane-induced lupus mouse model

The *Sting*^*gt/gt*^ mice (the goldenticket or *Tmem173*^*gt*^) mice were created via chemically inducing mutagen with N**-**ethyl**-**N**-**nitrosourea (ENU) (27). The C57BL/6 WT and *Sting*^*gt/gt*^ mice (8-10 weeks old) were intraperitoneally injected with 500 μl of pristane or tetramethylpentadecane (TMPD) (#P2870, SIGMA-ALDRICH Co., MO, USA) (4). All experiments were performed with the approval of the Animal Experimentation Ethics Committee of Chulalongkorn University Medical School with all relevant institutional guidelines.

### Autoantibody detection

The anti-dsDNA was performed from the sera (1;100) collected six months after pristane injection by ELISA (26). The anti-nuclear antibody (ANA) was detected in diluted sera (1:2000) by immunofluorescence technique (26).

### Flow cytometry

The isolated splenocytes were performed as described (26). The isolated splenocytes (1 × 10^6^ cells) were stained with antibodies, as mentioned in the **Supplementary material**.

### Immunohistochemistry

The mice were euthanized six months after pristane injection. The whole lung was infused with paraformaldehyde and immersed in 4% paraformaldehyde/PBS. The kidneys were fixed with 4% paraformaldehyde/PBS. The tissues were embedded in paraffin, sectioned, and stained with H&E. Tissues were blindly graded using glomerular and interstitial scores for kidneys and diffuse pulmonary hemorrhage and interstitial inflammation for lungs (30, 31).

### Study population

One hundred seventy-three healthy blood donors and 302 SLE patients followed up between 2016-2021 at Ramathibodi hospital were enrolled. The inclusion criteria are > 18 years and diagnosed SLE using the 1997 ACR criteria (32) or SLICC criteria 2012 (33) for SLE classification. The exclusion criteria are a history of cancer. The medical records were reviewed and analyzed if the patients had followed up for at least five years since the diagnosis. The study (MURA2015/731and MURA2021/177)) was approved by the Faculty of Medicine Ramathibodi Hospital ethics committee and conducted according to the guidelines of the Declaration of Helsinki.

### Identification of STING genotype in SLE patients

We designed TaqMan probes to detect STING variants at c.212G>A (R71H, rs11554776, Assay ID C_62979_10), c.689G>C (G230A, rs78233829, Assay ID C_104371077_10), c.695G>A (R232H, rs1131769, Assay ID C_26015196_10), c.878G>A (R293Q, rs7380824, Assay ID C_28947918_10), and mutation at c.852G>T were designed and customized by the author (R284S, Assay ID ANXGV44).

### Clinical assessment

Disease activity at diagnosis, 1^st^ and 5^th^ year of follow-up was assessed by systemic lupus erythematosus disease activity index 2000 (SLEDAI-2K) (34), and remission was defined as clinical SLEDAI = 0 and physician global assessment <0.5 by the 2021 DORIS definition of remission in SLE (35). Disease flare was defined as SLEDAI-2K increase ≥3 (34). End organ damage in 5^th^ year of follow-up as measured by Systemic Lupus International Collaborating Clinic/American College of Rheumatology Damage Index (36).

### Statistical analysis

The descriptive characteristics were presented by mean ± standard deviation and frequencies (%). The categorical variables were compared using Chi-squared or Fisher’s exact test. The continuous variables were compared using a t-test or Mann-Whitney U-test between two groups. Logistic regression and linear regression analysis were performed to identify the independent factors associated with inheritance patterns and the effect of the variants in SLE patients. All analyses were performed using STATA version 15 (Stat Corp., College Station, Texas, USA), and the statistical significance was defined as a p-value of <0.05.

## RESULTS

### *Sting/Tmem173* mediated autoantibody production in Pristane-induced lupus

Autoreactive B cells produced the anti-nuclear antibody (ANA), the autoantibody commonly found in lupus disease. Pristane induced autoantibody production, including anti-Sm, RNP, dsDNA, chromatin, and ribosomal P (37, 38). We tested for ANA from the sera collected six months after pristane injection to demonstrate whether STING affects autoantibody production. The pattern of ANA showed similarity in both C57/BL6-WT and *Sting*^*gt/gt*^ mice. However, the intensity of ANA staining identified by immunofluorescence was significantly reduced in *Sting*^*gt/gt*^ mice compared to C57/BL6-WT mice (Figures 1A-1B). Also, the level of anti-dsDNA titers did not increase by pristane injection in the *Sting*^*gt/gt*^ mice (Figure 1C).

**Figure 1.**
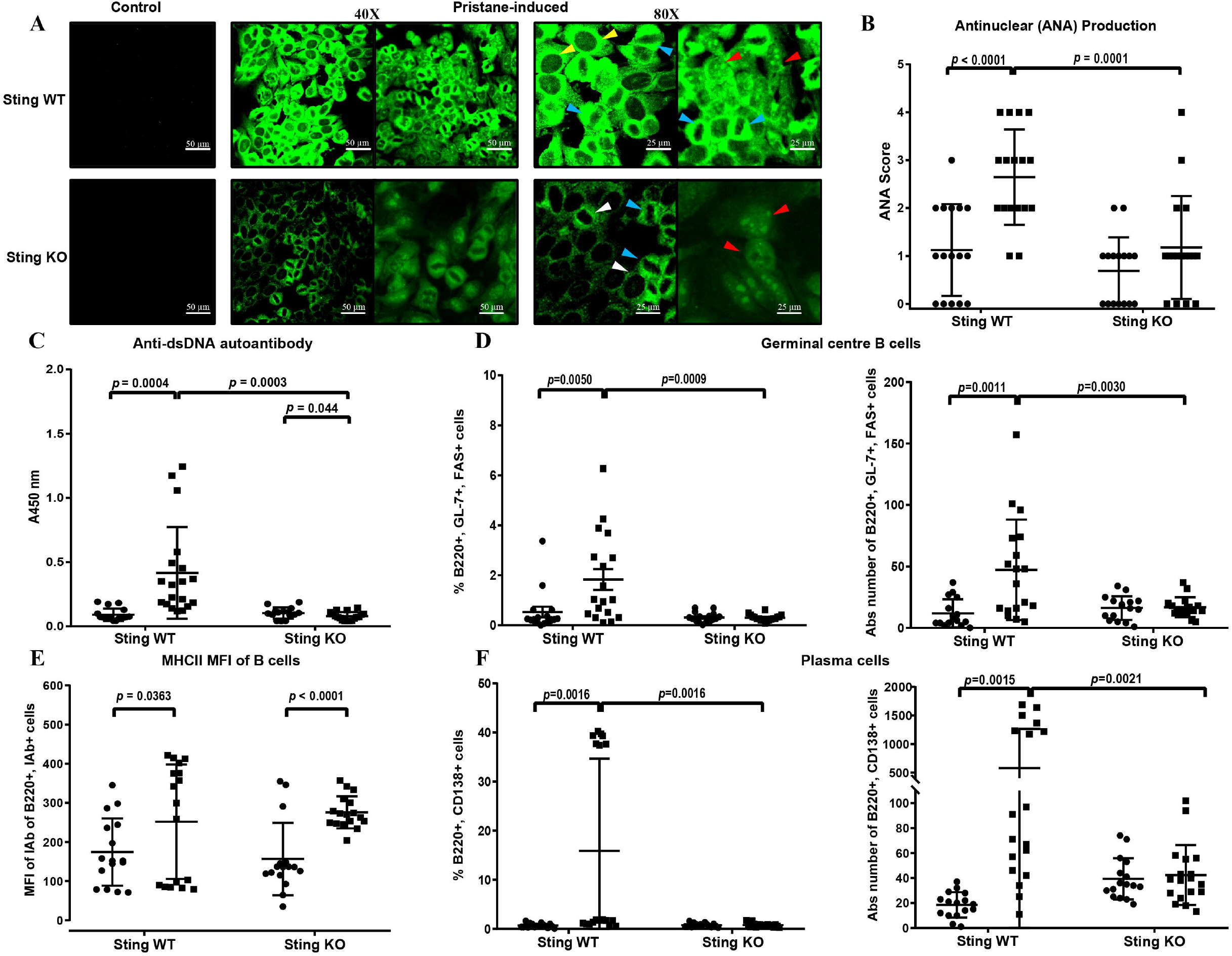
*Sting/Tmem173* mediated autoantibody production in Pristane-induced lupus. (A) Confocal microscopy shows staining of an anti-nuclear antibody (ANA) of Sting WT (*Sting*^*wt/wt*^) and Sting-deficient (*Sting*^*gt/gt*^) mouse sera at 1:2000 dilution. The ANA patterns show cytoplasmic homogenous (yellow arrow), mitotic (blue arrow), rim (white arrow), and nucleolar (red arrow). Scale bar 50 μM at 40X magnification and 25 μM at 80X magnification. (B) The fluorescence intensity of ANA was scored at the dilution of 1:2,000. (C) Detection of anti-double-stranded DNA (dsDNA) (1:100) using ELISA, read out at the wavelength of 450 nm. (D) Flow cytometry analysis of splenocytes shows the percentage and cell number of germinal center B cells (B220^+^GL-7^+^FAS^+^) and plasma cells (B220^+^CD138^+^). (E) The mean fluorescence intensity (MFI) of IA^b^ represents MHC II expression in the B cells. Data are shown as mean ± SEM; *p*-values indicated in the figures (N = 16-18 mice/group).

*Sting/Tmem173* is involved in spontaneous germinal center B cell development and plasma cell differentiation, requiring autoantibody production in the Fcgr2b-deficient mice (26). The expansion of germinal center B cells (Figure 1D) and plasma cells (Figure 1F) were absent in the *Sting*^*gt/gt*^ mice after pristane injection. However, Pristane induced the mean fluorescence intensity of MHC (IAb) on B cells representing the antigen-presenting ability, in a similar pattern in both WT and *Sting*^*gt/gt*^ B cells (Figure 1E).

The data suggested that STING is involved in autoantibody production, germinal center formation, and plasma cell expansion.

### *Sting/Tmem173* facilitated adaptive and innate immunity activation in Pristane-induced lupus

Dendritic cell (DC) expansion did not occur in the *Sting*^*gt/gt*^ mice after pristane injection (Figures 2A-2B). Although Pristane induced the activating classical DC (cDC) in comparable numbers in WT and *Sting*^*gt/gt*^ mice (Fig.2C-2D), the expansion of plasmacytoid dendritic cells (pDC) was absent in *Sting*^*gt/gt*^ mice but not WT mice (Figures 2E-2F). The induction of neutrophils increased in both WT and *Sting*^*gt/gt*^ ((Figures 2G-2H). The increase in the macrophage numbers only occurred in WT mice (Figures 2I-2J). Interestingly, both WT and *Sting*^*gt/gt*^ mice showed a rise in MHC expression in cDC and macrophages after pristane injection (Figures 2K-2L). The data suggested STING participated in pDC and macrophage expansion in pristane-induced lupus.

**Figure 2.**
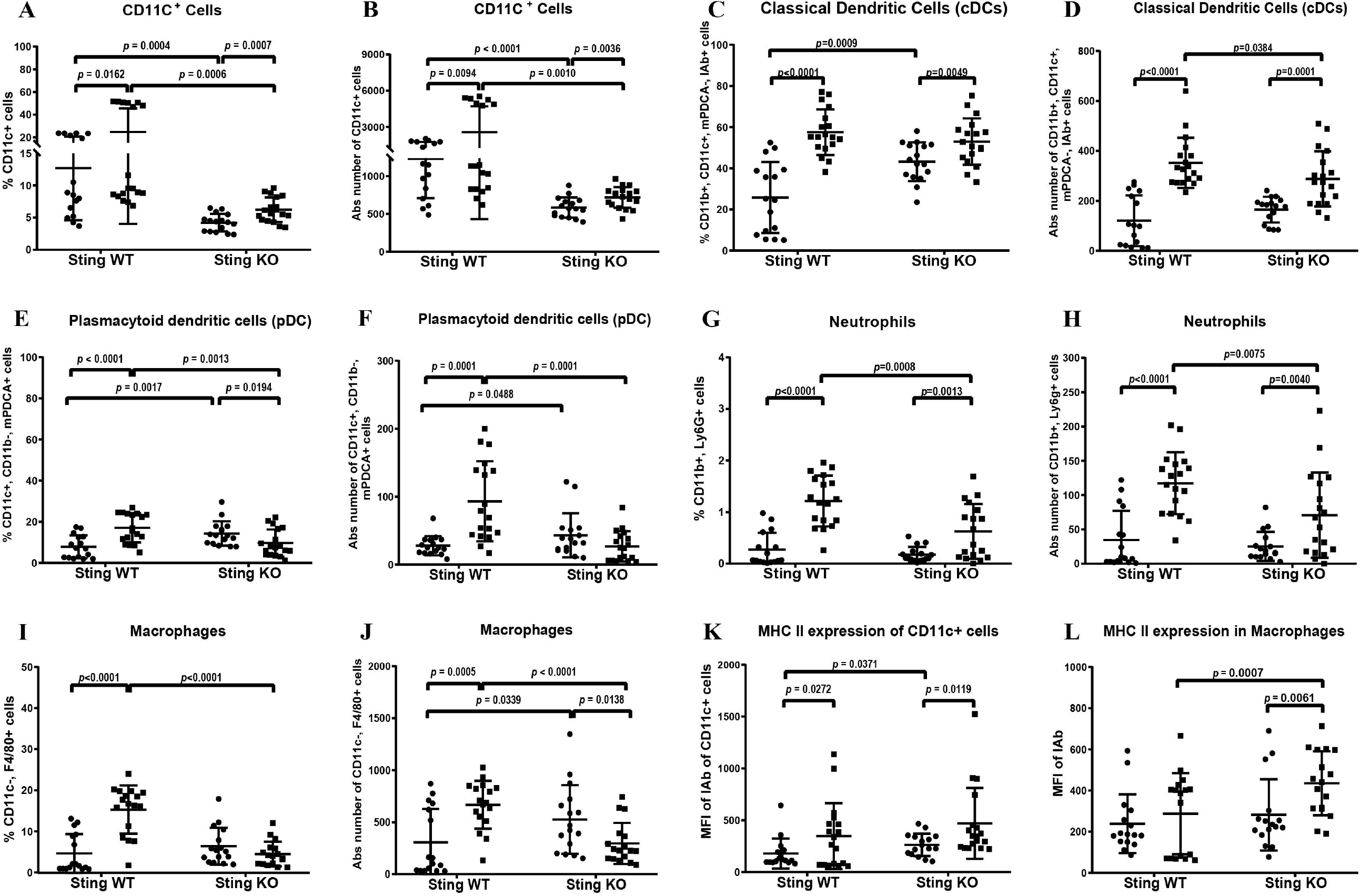
*Sting/Tmem173* facilitated plasmacytoid dendritic cells and macrophage expansion in Pristane-induced lupus. The isolated splenocytes were stained and analyzed by flow cytometry. Data show the proportion and the absolute number of cells (A–B) DC (CD11c^+^), (C – D) activated cDC (CD11c^+^CD11b^+^mPDCA^-^IAb^+^), (E – F) pDC (CD11c^+^CD11b^-^mPDCA^+^), (G – H) neutrophils (CD11b^+^Ly6g^+^), and (I – J) macrophages (CD11c^-^F480^+^). (K-L) The MFI of IAb^+^ represents MHC II expression of (K) DC and (L) macrophages. Data are shown as mean ± SEM; *p*-values indicated in the figures (N = 16-18 mice/group).

Activated DC primed naive T cells to become effector memory cells. The expansion of effector memory T cells in the *Fcgr2b*-deficient mice declined without STING signaling (26). However, the proportion of effector memory T cells in WT and Sting-gt increased in the pristane-induced lupus model (Supplementary Figure 1A). The IFN-γ-producing T helper cells expanded, but IL-17A-producing T helper cells decreased after pristane injection in the *Sting*^*gt/gt*^ mice (Supplementary Figures 1B-1C). STING antagonist reduced the double-negative (DN) T cells in the *Fcgr2b*-deficient mice (28). Similarly, pristane injection in the *Sting*^*gt/gt*^ mice did not increase the DN-T cells (Supplementary Figure 1D). DN-T cells produced both IFN-γ and IL-17A in lupus mice (39). Interestingly, IL-17A^+^ DN-T cells, not IFN-γ^+^ DN-T cells, were reduced in the *Sting*^*gt/gt*^ mice in pristane-induced lupus (Supplementary Figures 1E-1F).

### STING-mediated inflammatory pathology in Pristane-induced lupus

The glomerular showed increased cellularity and mesangial expansion in WT and the *Sting*^*gt/gt*^ mice (Figure 3A). However, the *Sting*^*gt/gt*^ mice developed lower glomerular and interstitial scores than WT mice after Pristane injection (Figures 3B-3C). The lungs of *Sting*^*gt/gt*^ mice significantly showed less inflammatory reaction compared to WT mice. The pathology showed diffuse pulmonary hemorrhage in WT mice but considerably decreased in the *Sting*^*gt/gt*^ mice (Figures 3D-3E). We did not detect the difference in the interstitial inflammation scores in WT and the *Sting*^*gt/gt*^ mice (Figure 3F); the severity in the WT mice showed mild infiltration. The data suggested that STING was responsible for the inflammatory response to Pristane-induced lupus.

**Figure 3.**
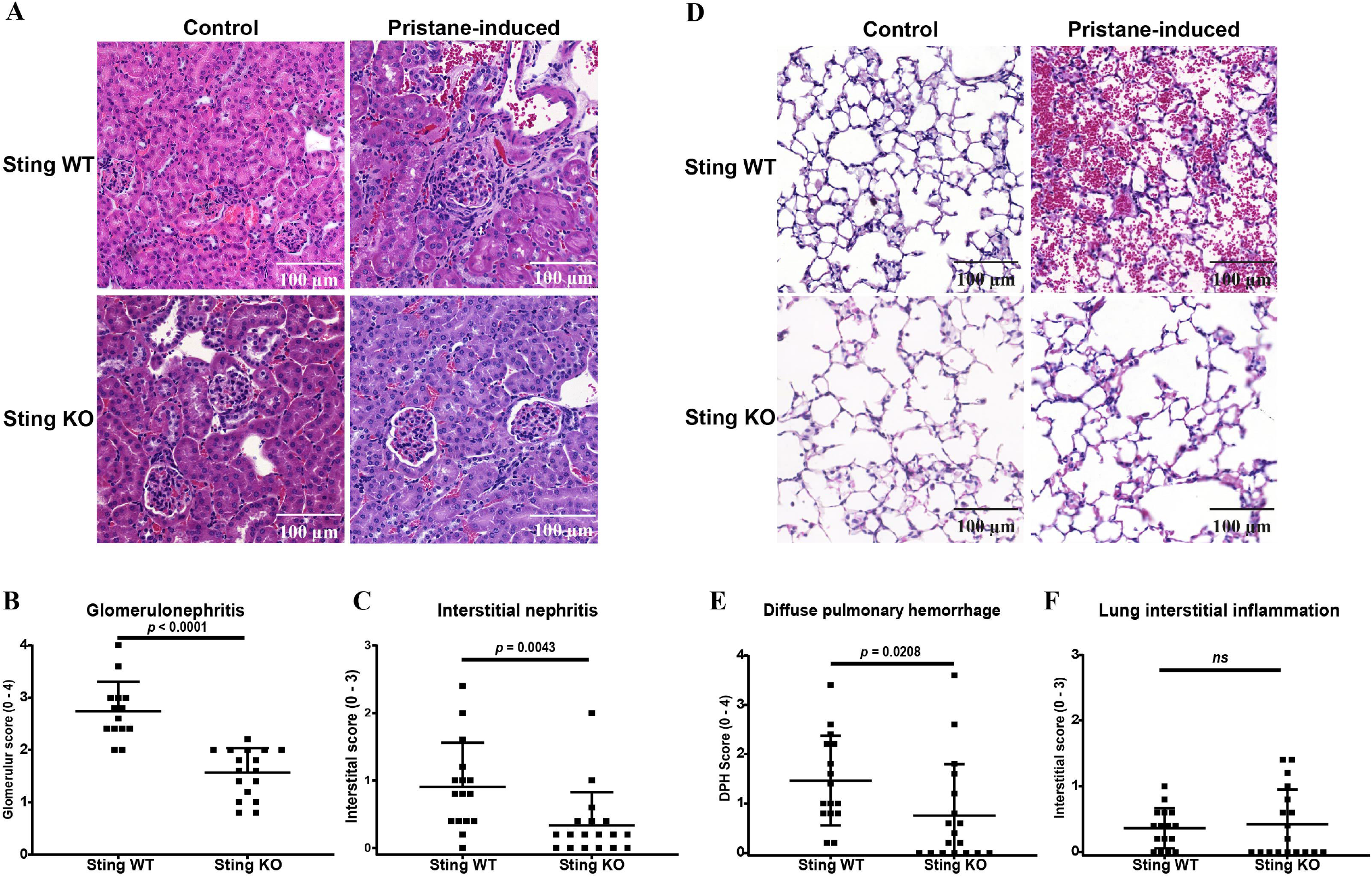
STING mediated inflammatory pathology in Pristane-induced lupus. The tissues (kidney and lung) were sectioned and stained with Hematoxylin and eosin stain (H&E). (A) The section of kidneys from pristane-induced mice showed glomerulonephritis (scale bar 100 μm). (B-C) The kidney histopathology was graded for (B) glomerulonephritis (glomerular score) and (C) interstitial nephritis (interstitial score). (D) The lung sections showed diffuse pulmonary hemorrhage from pristane-induced mice (scale bar 100 μm). (E-F) The lung histopathology was graded for (E) diffuse pulmonary hemorrhage and (F) interstitial inflammation scores. Data are shown as mean ± SEM; *p*-values indicated in the figures (N = 16-18 mice/group).

### STING variants and inheritance patterns in SLE patients

To explore the role of *STING* in human SLE, we decided to test the effect of *STING* variants on the genetic susceptibility of SLE. The SLE patients in this study were diagnosed at various ages (Supplementary Figure 2), not relatives. We tested the common *STING* single nucleotide polymorphisms (SNPs) that create missense mutation and have been reported in the general population as the followings: Rs11554776 (c.212G>A, R71H), Rs78233829 (c.689G>C, G230A), Rs7380824 (c.878G>A, R293Q), and Rs1131769 (c.695G>A, R232H) (40). To identify the risk of SLE patients with STING gain-of-function mutation, we examined the *STING* mutant (c.852G>T, R284S), which did not require cyclic dinucleotides to increase activity and was reported in STING-induced inflammatory disease (41).

We identified the inheritance pattern of each variant. Three hundred two patients with SLE and 173 healthy donors were tested for STING SNPs and mutation. The results showed that G230A and R284S increased OR with autosomal dominant inheritance pattern while R71H and R293Q with autosomal recessive pattern increased the OR for SLE (Table 1). Interestingly, the gain-of-function mutation at R284S is the highest risk allele [OR = 64.086 (95%CI 22.8605 –179.6555), p <0.0001]. Interestingly, R232H was described as a loss-of-function *Sting* mutation (42) decreased the OR of SLE [OR = 0.2515 (95%CI = 0.1648-0.3836), p < 0.0001].

**Table 1.** *STING* variants and inheritance patterns in SLE patients

| Rs number | variants | Inheritance pattern |  |  |  |  |  |
| --- | --- | --- | --- | --- | --- | --- | --- |
|  |  | Autosomal dominant |  |  | Autosomal recessive |  |  |
|  |  | OR | 95%CI | p-value | OR | 95%CI | p-value |
| Rs11554776 | c.212G>A<br>*R71H* | 1.0962 | 0.7494-1.6036 | 0.636 | 1.5782 | 1.0537-2.3636 | 0.027 |
| Rs78233829 | c.689G>C<br>*G230A* | 3.8474 | 2.3973-6.1747 | <0.0001 | 4.0107 | 2.4409-6.5898 | <0.0001 |
| Rs7380824 | c.878G>A<br>*R293Q* | 0.9475 | 0.6423-1.3976 | 0.786 | 2.5940 | 1.5780-2.4642 | <0.0001 |
| Rs1131769 | C.695G>A<br>*R232H* | 0.8236 | 0.3913-1.7335 | 0.609 | 0.2515 | 0.1648-0.3836 | <0.0001 |
| N/A | c.852G>T<br>*R284S* | 64.0860 | 22.8605-179.6555 | <0.0001 | N/A | N/A | N/A |

### The genotype pattern of *STING* variants in SLE susceptibility

Our analysis showed that *R71H* and *R293Q* in autorecessive inheritance and *G230A* in autosomal dominant increased the OR for SLE (Table 1). Thus, we defined the HAQ alleles in this study as H^AR^A^AD^Q^AR^ and divided them into complete-*HAQ* alleles, incomplete-*HAQ* alleles, and absence-HAQ alleles. The complete-HAQ alleles mean substitution in all three variants with the *H*^*AR*^*A*^*AD*^*Q*^*AR*^ pattern, and incomplete-*HAQ* alleles mean substitution in at least one but not all three variants. If patients have no substitution, we defined it as the absence-*HAQ* allele. The complete -HAQ group increased the SLE risk [OR=2.7256 (95%CI 1.5838-4.6905), p < 0.0001]. The absent-*HAQ* group decreased the OR for SLE [OR=0.3583 (95%CI=0.2166-0.5926), p < 0.0001], while the incomplete-HAQ did not increase the SLE risk **(Table2)**. The result suggested the additional effect of HAQ alleles in specific inheritance patterns increases SLE risk in this population.

**Table 2.** The genotype pattern of *STING* in SLE susceptibility

| Genotype pattern | Healthy | SLE | OR | 95%CI | P-value |
| --- | --- | --- | --- | --- | --- |
| Complete-HAQ | 19 | 76 | 2.7256 | 1.5838-4.6905 | <0.0001 |
| Incomplete-HAQ | 111 | 194 | 1.0033 | 0.6794-1.4815 | 0.987 |
| Absent-HAQ | 43 | 32 | 0.3583 | 0.2166-0.5926 | <0.0001 |
| Complete HAQ w/o S284 | 12 | 1 | N/A | N/A | N/A |
| Complete HAQ w/S284 | 7 | 75 | 128.5714 | 14.5033-1139.778 | <0.0001 |
| Complete HAQ w/o H232 | 3 | 41 | N/A | N/A | N/A |
| Complete HAQ w/ H232 | 16 | 35 | 0.16 | 0.0430-0.5950 | 0.006 |

The mutation at *R284S* is a gain-of-function of STING, which increases IFN-I in the absence of ligands (43), and dramatically enhances the SLE risk when detected in the complete-HAQ background [OR=128.5714 (95%CI 14.5033-1139.778), p-value< 0.0001] **(Table2)**. Interestingly, the loss of function STING mutation (R232H) significantly decreased SLE risk in the complete-HAQ susceptibility background [OR = 0.16 (95%CI 0.0430-0.5950), p-value 0.006) **(Table 2)**. The data suggested the complex interaction of STING genes in SLE susceptibility and the synergistic effect of multiple *STING* alleles on human SLE susceptibility.

### Comparison of clinical manifestations among *STING* genotype patterns in SLE patients

Among 302 patients with *STING* genotypes, only 201 patients showed complete medical records for the analysis (Supplementary Figure 2). The distributions of HAQ alleles, R284S, and R232H, were shown (Supplementary Figure 2). Almost all of these SLE patients showed R284S mutation (199 out of 201). Thus, we compared the clinical manifestation between complete-HAQ and non-complete-HAQ on the R284S background and did not see a difference in clinical parameters (Table 3). The data suggested that R284S, the gain-of-function allele, is the dominant variant causing the SLE phenotypes over the HAQ alleles. Furthermore, we analyzed the effect of R232H, the loss-of-function allele, on the non-complete-HAQ with R284S background and identified the lower SLE disease activity at the time of diagnosis (Table 3). These data suggested that SLE patients with non-risk alleles of *STING* could benefit from the loss-of-function mutation to present with low disease activity at diagnosis.

**Table 3.** Comparison of clinical manifestations among *STING* genotype patterns in SLE patients

| Clinical Parameters | Total SLE<br>(N = 201) | R284S w/o R232H<br>(N = 104) |  | P-value | Non-complete HAQ w/R284S<br>(N = 149) |  | P-value |
| --- | --- | --- | --- | --- | --- | --- | --- |
|  |  | Complete HAQ<br>(N=26) | Non-complete<br>HAQ (N=78) |  | W/R232H<br>(N=71) | W/O R232H<br>(N=78) |  |
| Age of onset (Mean±SD) | 29.67±10.60 | 31.96±11.95 | 29.25±8.81 | 0.219 | 29.49, 11.69 | 29.25, 8.81 | 0.888 |
| Female | 192 (95.52) | 25 (96.15) | 74 (94.87) | 0.791 | 68 (95.77) | 74 (94.87) | 1.000 |
| Clinical manifestations |  |  |  |  |  |  |  |
| Skin involvement | 181 (90.05) | 25 (96.15) | 72 (92.31) | 0.498 | 61 (85.92) | 72 (92.31) | 0.208 |
| MSK involvement | 150 (74.63) | 17 (65.38) | 57 (73.08) | 0.453 | 55 (77.46) | 57 (73.08) | 0.536 |
| Renal involvement | 149 (74.63) | 20 (76.92) | 55 (70.51) | 0.528 | 56 (78.87) | 55 (70.51) | 0.242 |
| Hematological involvement | 145 (72.14) | 19 (73.08) | 56 (71.79) | 0.900 | 51 (71.83) | 56 (71.79) | 0.996 |
| Cardiopulmonary involvement | 30 (14.93) | 6 (23.08) | 9 (11.54) | 0.196 | 12 (16.90) | 9 (11.54) | 0.347 |
| Neurological involvement | 46 (22.89) | 7 (26.92) | 19 (24.36) | 0.794 | 16 (22.54) | 19 (24.36) | 0.793 |
| GI involvement | 16 (7.96) | 4 (15.38) | 5 (6.41) | 0.223 | 6 (8.45) | 5 (6.41) | 0.634 |
| Constitutional symptoms | 62 (30.85) | 7 (26.92) | 25 (32.05) | 0.624 | 19 (26.76) | 25 (32.05) | 0.480 |
| anti-sm | 36 (17.91) | 8 (30.77) | 15 (19.23) | 0.220 | 11 (15.49) | 15 (19.23) | 0.548 |
| anti-dsDNA | 111 (55.22) | 16 (61.54) | 42 (53.85) | 0.494 | 42 (59.15) | 42 (53.85) | 0.514 |
| SLEDAI score at diagnosis (Mean±SD) | 12.12±6.93 | 11.92±6.58 | 13.26±6.86 | 0.384 | 11.49±7.40 | 13.26±6.86 | 0.131 |
| Mild and moderate activity | 119 (59.2) | 15 (57.69) | 39 (50.00) | 0.497 | 49 (69.01) | 39 (50.00) | 0.018* |
| Severe activity | 82 (40.80) | 11 (42.31) | 39 (50.00) |  | 22 (30.99) | 39 (50.00) |  |
| Remission at 1<sup>st</sup> year | 56 (26.86) | 8 (30.77) | 20 (25.64) | 0.61 | 18 (25.35) | 20 (25.64) | 0.968 |
| Remission at 5<sup>th</sup> year | 89 (44.28) | 14 (53.85) | 33 (42.31) | 0.61 | 33 (46.48) | 33 (42.31) | 0.609 |
| Disease flare ≥ 2 times | 22 (10.95) | 5 (19.23) | 6 (7.69) | 0.137 | 7 (9.86) | 6 (7.69) | 0.64 |
| Free of steroids in 5<sup>th</sup> year | 44 (21.89) | 6 (23.08) | 15 (19.23) | 0.672 | 18 (25.35) | 15 (19.23) | 0.369 |
| Free of immunosuppressive at 5<sup>th</sup> year | 29 (14.5) | 7 (26.92) | 11 (14.10) | 0.145 | 9 (12.68) | 11 (14.10) | 0.799 |
| SLICC/ACR damage score ≥ 1 in 5<sup>th</sup> year | 54 (26.87) | 5 (19.23) | 24 (30.77) | 0.256 | 14 (19.72) | 24 (30.77) | 0.122 |
| Death | 4 (1.99%) | 1 (3.85) | 0 (0) | 0.25 | 1 (1.41) | 0 | 0.477 |

Definition: non-complete *HAQ* = both incomplete and absence *HAQ*

## DISCUSSION

The background of the lupus mouse models suggested the different mechanisms involved in disease development (4). The MRL.*Fas*^*lpr*^ mice, which do not have IFN signatures, require STING-mediated signaling, but the disease aggravates in the absence of STING (4, 24). In contrast, the lack of STING signaling rescues the *Trex1*-deficient mice, requiring type I IFN-mediated signaling (11, 14, 15). The 129/B6.*Fcgr2b-*deficient mice contained 129 loci autoimmune susceptibility increased IFN signature and survived without STING mediated pathway (26). The enrichment of type I IFN signature in the lupus mouse might indicate the role of STING in disease development in a specific model.

We detected the decrease of ANA, anti-dsDNA together with germinal center B cells, and plasma cells in the Sting-deficient on Goldenticket mutant mice, similar findings reported in the 129/B6.*Fcgr2b*-deficient mice (26). The *Sting*^*gt/gt*^ mice reduced the expansion of pDC, and the numbers of neutrophils and macrophages after pristane injection, while the conventional dendritic cells did not change. The differentiation of pDC is STING-dependent (26). However, the mean fluorescence intensity of MHC-II on macrophages was higher in the *Sting*^*gt/gt*^ mice, which suggested the phenotype of activated macrophages was STING-independent. These data suggested the diverse role of STING signaling on individual cell types, which might affect organ-specific phenotypes.

The activated cDC promotes Tem differentiation in the 129/B6.*Fcgr2b*-deficient mice (26). The expansion of cDC was STING-dependent in the 129/B6.*Fcgr2b*-deficient mice but not in pristane-induced lupus. In contrast to the 129/B6.*Fcgr2b-*deficient mice whose T effector memory cells (Tem) were reduced in the absence of STING, the pristane-induced lupus model induced the Tem in both the *Sting*^*gt/gt*^ and WT mice in a similar manner. The difference in Tem phenotypes between these two models may derive from the ability of activated cDC to prime T cells. Lupus mouse models and human SLE studies show the increase of double-negative T cells (DN-T), which produced both IFN-γ and IL-17A (39, 44). Blocking the IL-17 signaling pathway ameliorates glomerulonephritis (39). Only IL-17^+^CD4^+^ cells but not IFN-γ^+^ CD4^+^ cells decreased in the *Sting*^*gt/gt*^ mice after pristane injection. Reducing IL-17-producing cells could be one of the mechanisms that reduce lupus nephritis severity.

In this study, we induced the lupus phenotypes (LN and DPH) in WT mice, allowing us to compare STING-deficient’s effect on Goldenticket mutant mice (*Sting*^*gt/gt*^) on lupus development. The severity of lupus nephritis and diffuse pulmonary hemorrhage in the *Sting*^*gt/gt*^ mice reduced significantly compared to WT after pristane injection. The findings suggested STING participated in the pathogenesis of pristane-induced lupus in the *Sting*^*gt/gt*^ mice. In contrast, a recent study shows that STING put a break on pristane-induced lupus mediated through endosomal Toll-like receptors. The *Sting*-deficient mice increased anti-nuclear antibodies, peritoneal macrophages, and CD11b^+^ Ly6C^hi^ inflammatory monocytes in the spleen. However, Pristane could not induce lupus nephritis in the WT mice in that study (45). The difference in findings in these two studies could be the backgrounds of the Sting-deficient mice. The *Sting*^*gt/gt*^ was created by the ENU mutagen (27), which could induce other mutations around the *Sting* locus and might influence lupus pathogenesis. The mice used in the previous study were *Sting* deletion (46). The mouse models that show the high activity of type I IFN required a STING signaling pathway (26, 47) while the MRL.Fas^l*pr*^ mice, mediated through endosomal TLRs, show the opposite result (24, 48).

In addition, a previous study did not induce lupus phenotypes (LN and DPH) after pristane injection (45). The lack of specific lupus pathology may suggest low expression of type I IFN in that environment, affecting the STING signaling mechanisms. Hence, it is difficult to justify whether the interruption of STING signaling did not affect lupus. Since the STING signaling pathway has demonstrated various functions in different cell types, models, and backgrounds, the evaluation of immunological markers without the disease-specific pathology may not draw the precise conclusion of pathogenesis overall. The complexity of lupus mouse models leads to the diverse mechanisms of SLE. *STING* mutations have been identified in human autoinflammatory disease and interferonpathies (18, 21). The specific STING inhibitor (ISD017) can ameliorate glomerulonephritis in the *Fcgr2b*-deficient mice and reduce ISG responses in peripheral blood mononuclear cells (PBMC) from SLE patients (28). The data suggested the intrinsic mechanism of STING underlying human SLE. Thus, we identified the STING role in human SLE by looking at the SNPs and mutation of *STING* in our SLE patients.

A previous study on multiple autoimmune diseases to identify STING SNP on exon five (Rs7380272) did not show the difference between patients and controls (23), which could be due to the small sample size or other *STING* variants associated with SLE susceptibility. We hypothesized that individual SNPs of STING’s effect on SLE risk would be minor, and the combinations of these SNPs might yield greater risk.

The *R71H, G230A*, and *R293Q* variants are HAQ alleles and could be found in up to 20% of the population (49). Although the *HAQ* homozygous allele was described as a loss-of-function mutation resulting in defective stimulation of type I IFN (50), another study showed that HAQ STING homozygous allele HAQ is indeed functionally responsive to cyclic dinucleotide (51). The *R71H* and R293Q variants cause the type I IFN-stimulating defect of *HAQ*, while the *G230A* variant was only a gain-of-function mutation in STING and resulted in an increased type I IFN (52). Here, we detected the inheritance pattern of each variant from SLE patients who showed a significant OR and combined the inheritance pattern of each variant (H^AR^ A^AD^ Q^AR^), considerably increasing the OR. The combination of 284S (gain-of-function) on top of the H^AR^A^AD^Q^AR^ inheritance pattern dramatically augmented SLE risk in this Thai population. Also, the loss-of-function STING variant (232H) reduced the SLE risk and disease severity at the time of diagnosis. This analysis revealed the risk of SLE development could derive from a perfect combination of *STING* variants providing synergistic effects. Our study showed that all SLE patients had at least one mutation, and almost all of the patients had a gain-of-function mutation, the R284S variant. This finding suggests that STING mutation may play an essential role in lupus development and may be a new target for SLE treatment.

## CONCLUSION

Our mouse and human SLE data suggested that STING-mediated signaling showed promising targeted intervention. Although some mouse models showed unencouraging results in inhibiting STING for lupus treatment, the difference in the results may depend on the background of the mice and intrinsic mechanisms that promote type I IFN production. Human SLE has high heterogeneity in disease phenotypes, severity, and genetic susceptibility. Since we identified that *STING* variants and mutation increased SLE risk, *STING* genetics could be a biomarker to identify patients who will respond to STING targeted treatment.

## Data Availability

Data are available upon reasonable request.

## AUTHOR CONTRIBUTIONS

PV, PP, TP, NH, and SP conceived the concept and design of this project. PV and PP designed the experiments. PP and TP supervised the experiments. PV and CT performed the experiments. PP, SW, and PN recruited patients. TS collected clinical samples. PV, CT, and SW collected demographic data from patients. AL, PV, and PP scored the pathological status of tissues. PV, SW, PN, and PP analyzed the data. PV and PP wrote and finalized the manuscript. All the authors reviewed, contributed, and approved this submitted manuscript.

## ACKNOWLEDGEMENTS

We thank all the great supporters who helped and facilitated us in performing this research. We appreciate our team members, Arthid Thim-uam and Mookmanee Tansakul, for helpful experimental suggestions and discussions, and the CUSB team members, especially Phitchapha Proykhunthod, Isara Alee, and Papasara Chantawichitwong, for your hands. Thanks to Meng-Shin Shiao, Saisuda Noojaroen, and Pamorn Chittavanich for arranging the facility to perform experiments.

## Ethics approval

All mouse experiments were performed with the approval of the Animal Experimentation Ethics Committee of Chulalongkorn University Medical School with all relevant institutional guidelines (007/2561). The SLE study (COA. MURA2015/731) was approved by the Faculty of Medicine Ramathibodi Hospital ethics committee and conducted according to the guidelines of the Declaration of Helsinki.

## Data availability

Data are available upon reasonable request.

## References

1. Dall’Era M, Cisternas MG, Snipes K, Herrinton LJ, Gordon C, Helmick CG. The Incidence and Prevalence of Systemic Lupus Erythematosus in San Francisco County, California: The California Lupus Surveillance Project. Arthritis Rheumatol. 2017;69(10):1996–2005.

2. Lewis MJ, Jawad AS. The effect of ethnicity and genetic ancestry on the epidemiology, clinical features and outcome of systemic lupus erythematosus. Rheumatology (Oxford). 2017;56(suppl_1):i67–i77.

3. Tsokos GC. Systemic lupus erythematosus. N Engl J Med. 2011;365(22):2110–21.

4. Zhuang H, Szeto C, Han S, Yang L, Reeves WH. Animal Models of Interferon Signature Positive Lupus. Front Immunol. 2015;6:291.

5. Santiago-Raber ML, Baccala R, Haraldsson KM, Choubey D, Stewart TA, Kono DH, et al. Type-I interferon receptor deficiency reduces lupus-like disease in NZB mice. J Exp Med. 2003;197(6):777–88.

6. Nacionales DC, Kelly-Scumpia KM, Lee PY, Weinstein JS, Lyons R, Sobel E, et al. Deficiency of the type I interferon receptor protects mice from experimental lupus. Arthritis Rheum. 2007;56(11):3770–83.

7. Pisitkun P, Deane JA, Difilippantonio MJ, Tarasenko T, Satterthwaite AB, Bolland S. Autoreactive B cell responses to RNA-related antigens due to TLR7 gene duplication. Science. 2006;312(5780):1669–72.

8. Deane JA, Pisitkun P, Barrett RS, Feigenbaum L, Town T, Ward JM, et al. Control of toll-like receptor 7 expression is essential to restrict autoimmunity and dendritic cell proliferation. Immunity. 2007;27(5):801–10.

9. Barrat FJ, Elkon KB, Fitzgerald KA. Importance of Nucleic Acid Recognition in Inflammation and Autoimmunity. Annu Rev Med. 2016;67:323–36.

10. Yang YG, Lindahl T, Barnes DE. Trex1 exonuclease degrades ssDNA to prevent chronic checkpoint activation and autoimmune disease. Cell. 2007;131(5):873–86.

11. Stetson DB, Ko JS, Heidmann T, Medzhitov R. Trex1 prevents cell-intrinsic initiation of autoimmunity. Cell. 2008;134(4):587–98.

12. Rice G, Newman WG, Dean J, Patrick T, Parmar R, Flintoff K, et al. Heterozygous mutations in TREX1 cause familial chilblain lupus and dominant Aicardi-Goutieres syndrome. Am J Hum Genet. 2007;80(4):811–5.

13. Crow YJ, Rehwinkel J. Aicardi-Goutieres syndrome and related phenotypes: linking nucleic acid metabolism with autoimmunity. Hum Mol Genet. 2009;18(R2):R130–6.

14. Gall A, Treuting P, Elkon KB, Loo YM, Gale M, Jr., Barber GN, et al. Autoimmunity initiates in nonhematopoietic cells and progresses via lymphocytes in an interferon-dependent autoimmune disease. Immunity. 2012;36(1):120–31.

15. Gray EE, Treuting PM, Woodward JJ, Stetson DB. Cutting Edge: cGAS Is Required for Lethal Autoimmune Disease in the Trex1-Deficient Mouse Model of Aicardi-Goutieres Syndrome. J Immunol. 2015;195(5):1939–43.

16. Summers SA, Hoi A, Steinmetz OM, O’Sullivan KM, Ooi JD, Odobasic D, et al. TLR9 and TLR4 are required for the development of autoimmunity and lupus nephritis in pristane nephropathy. J Autoimmun. 2010;35(4):291–8.

17. Bossaller L, Christ A, Pelka K, Nundel K, Chiang PI, Pang C, et al. TLR9 Deficiency Leads to Accelerated Renal Disease and Myeloid Lineage Abnormalities in Pristane-Induced Murine Lupus. J Immunol. 2016;197(4):1044–53.

18. Liu Y, Jesus AA, Marrero B, Yang D, Ramsey SE, Sanchez GAM, et al. Activated STING in a vascular and pulmonary syndrome. N Engl J Med. 2014;371(6):507–18.

19. d’Angelo DM, Di Filippo P, Breda L, Chiarelli F. Type I Interferonopathies in Children: An Overview. Front Pediatr. 2021;9:631329.

20. Keskitalo S, Haapaniemi E, Einarsdottir E, Rajamaki K, Heikkila H, Ilander M, et al. Novel TMEM173 Mutation and the Role of Disease Modifying Alleles. Front Immunol. 2019;10:2770.

21. Jeremiah N, Neven B, Gentili M, Callebaut I, Maschalidi S, Stolzenberg MC, et al. Inherited STING-activating mutation underlies a familial inflammatory syndrome with lupus-like manifestations. J Clin Invest. 2014;124(12):5516–20.

22. Konig N, Fiehn C, Wolf C, Schuster M, Cura Costa E, Tungler V, et al. Familial chilblain lupus due to a gain-of-function mutation in STING. Ann Rheum Dis. 2017;76(2):468–72.

23. Balada E, Selva-O’Callaghan A, Felip L, Ordi-Ros J, Simeon-Aznar CP, Solans-Laque R, et al. Sequence analysis of TMEM173 exon 5 in patients with systemic autoimmune diseases. Autoimmunity. 2016;49(1):12–6.

24. Sharma S, Campbell AM, Chan J, Schattgen SA, Orlowski GM, Nayar R, et al. Suppression of systemic autoimmunity by the innate immune adaptor STING. Proc Natl Acad Sci U S A. 2015;112(7):E710–7.

25. Tansakul M, Thim-Uam A, Saethang T, Makjaroen J, Wongprom B, Pisitkun T, et al. Deficiency of STING Promotes Collagen-Specific Antibody Production and B Cell Survival in Collagen-Induced Arthritis. Front Immunol. 2020;11:1101.

26. Thim-Uam A, Prabakaran T, Tansakul M, Makjaroen J, Wongkongkathep P, Chantaravisoot N, et al. STING Mediates Lupus via the Activation of Conventional Dendritic Cell Maturation and Plasmacytoid Dendritic Cell Differentiation. iScience. 2020;23(9):101530.

27. Sauer JD, Sotelo-Troha K, von Moltke J, Monroe KM, Rae CS, Brubaker SW, et al. The N-ethyl-N-nitrosourea-induced Goldenticket mouse mutant reveals an essential function of Sting in the in vivo interferon response to Listeria monocytogenes and cyclic dinucleotides. Infect Immun. 2011;79(2):688–94.

28. Prabakaran T, Troldborg A, Kumpunya S, Alee I, Marinkovic E, Windross SJ, et al. A STING antagonist modulating the interaction with STIM1 blocks ER-to-Golgi trafficking and inhibits lupus pathology. EBioMedicine. 2021;66:103314.

29. Kennedy RB, Haralambieva IH, Ovsyannikova IG, Voigt EA, Larrabee BR, Schaid DJ, et al. Polymorphisms in STING Affect Human Innate Immune Responses to Poxviruses. Front Immunol. 2020;11:567348.

30. Shi Y, Tsuboi N, Furuhashi K, Du Q, Horinouchi A, Maeda K, et al. Pristane-induced granulocyte recruitment promotes phenotypic conversion of macrophages and protects against diffuse pulmonary hemorrhage in Mac-1 deficiency. J Immunol. 2014;193(10):5129–39.

31. Wu EK, Henkes ZI, McGowan B, Bell RD, Velez MJ, Livingstone AM, et al. TNF-Induced Interstitial Lung Disease in a Murine Arthritis Model: Accumulation of Activated Monocytes, Conventional Dendritic Cells, and CD21(+)/CD23(-) B Cell Follicles Is Prevented with Anti-TNF Therapy. J Immunol. 2019;203(11):2837–49.

32. Hochberg MC. Updating the American College of Rheumatology revised criteria for the classification of systemic lupus erythematosus. Arthritis Rheum. 1997;40(9):1725.

33. Petri M, Orbai AM, Alarcon GS, Gordon C, Merrill JT, Fortin PR, et al. Derivation and validation of the Systemic Lupus International Collaborating Clinics classification criteria for systemic lupus erythematosus. Arthritis Rheum. 2012;64(8):2677–86.

34. Gladman DD, Ibañez D, Urowitz MB. Systemic lupus erythematosus disease activity index 2000. J Rheumatol. 2002;29(2):288–91.

35. van Vollenhoven RF, Bertsias G, Doria A, Isenberg D, Morand E, Petri MA, et al. 2021 DORIS definition of remission in SLE: final recommendations from an international task force. Lupus Sci Med. 2021;8(1).

36. Gladman D, Ginzler E, Goldsmith C, Fortin P, Liang M, Urowitz M, et al. The development and initial validation of the Systemic Lupus International Collaborating Clinics/American College of Rheumatology damage index for systemic lupus erythematosus. Arthritis Rheum. 1996;39(3):363–9.

37. Satoh M, Kumar A, Kanwar YS, Reeves WH. Anti-nuclear antibody production and immune-complex glomerulonephritis in BALB/c mice treated with pristane. Proc Natl Acad Sci U S A. 1995;92(24):10934–8.

38. Satoh M, Reeves WH. Induction of lupus-associated autoantibodies in BALB/c mice by intraperitoneal injection of pristane. J Exp Med. 1994;180(6):2341–6.

39. Pisitkun P, Ha HL, Wang H, Claudio E, Tivy CC, Zhou H, et al. Interleukin-17 cytokines are critical in development of fatal lupus glomerulonephritis. Immunity. 2012;37(6):1104–15.

40. Patel S, Jin L. TMEM173 variants and potential importance to human biology and disease. Genes Immun. 2019;20(1):82–9.

41. Konno H, Chinn IK, Hong D, Orange JS, Lupski JR, Mendoza A, et al. Pro-inflammation Associated with a Gain-of-Function Mutation (R284S) in the Innate Immune Sensor STING. Cell Rep. 2018;23(4):1112–23.

42. Walker MM, Kim S, Crisler WJ, Nguyen K, Lenz LL, Cambier JC, et al. Selective Loss of Responsiveness to Exogenous but Not Endogenous Cyclic-Dinucleotides in Mice Expressing STING-R231H. Front Immunol. 2020;11:238.

43. Melki I, Rose Y, Uggenti C, Van Eyck L, Fremond ML, Kitabayashi N, et al. Disease-associated mutations identify a novel region in human STING necessary for the control of type I interferon signaling. J Allergy Clin Immunol. 2017;140(2):543–52 e5.

44. Rafael-Vidal C, Perez N, Altabas I, Garcia S, Pego-Reigosa JM. Blocking IL-17: A Promising Strategy in the Treatment of Systemic Rheumatic Diseases. Int J Mol Sci. 2020;21(19).

45. Motwani M, McGowan J, Antonovitch J, Gao KM, Jiang Z, Sharma S, et al. cGAS-STING Pathway Does Not Promote Autoimmunity in Murine Models of SLE. Front Immunol. 2021;12:605930.

46. Ishikawa H, Barber GN. STING is an endoplasmic reticulum adaptor that facilitates innate immune signalling. Nature. 2008;455(7213):674–8.

47. Ahn J, Gutman D, Saijo S, Barber GN. STING manifests self DNA-dependent inflammatory disease. Proc Natl Acad Sci U S A. 2012;109(47):19386–91.

48. Christensen SR, Shupe J, Nickerson K, Kashgarian M, Flavell RA, Shlomchik MJ. Toll-like receptor 7 and TLR9 dictate autoantibody specificity and have opposing inflammatory and regulatory roles in a murine model of lupus. Immunity. 2006;25(3):417–28.

49. Yi G, Brendel VP, Shu C, Li P, Palanathan S, Cheng Kao C. Single nucleotide polymorphisms of human STING can affect innate immune response to cyclic dinucleotides. PLoS One. 2013;8(10):e77846.

50. Patel S, Blaauboer SM, Tucker HR, Mansouri S, Ruiz-Moreno JS, Hamann L, et al. The Common R71H-G230A-R293Q Human TMEM173 Is a Null Allele. J Immunol. 2017;198(2):776–87.

51. Sivick KE, Surh NH, Desbien AL, Grewal EP, Katibah GE, McWhirter SM, et al. Comment on “The Common R71H-G230A-R293Q Human TMEM173 Is a Null Allele”. J Immunol. 2017;198(11):4183–5.

52. Jin L, Xu LG, Yang IV, Davidson EJ, Schwartz DA, Wurfel MM, et al. Identification and characterization of a loss-of-function human MPYS variant. Genes Immun. 2011;12(4):263–9.

