## Supplementary Data for "Role of *STING/TMEM173* mutation in systemic lupus erythematosus: from animal model to intrinsic human genetics"

**SUPPLEMENTARY DOCUMENT**

**SUPPLEMENTARY MATERIALS AND METHODS**

**Animals**

The *Sting*$^{gt/gt}$ mice were obtained from Professor Paludan (Aarhus University, Denmark), while wild-type (C57BL/6) mice were purchased from the National Laboratory Animal Center, Nakornpathom, Thailand. The *Sting*$^{gt/gt}$ mice (the goldenticket or *Tmem173*$^{gt}$) mice were created via chemically inducing mutagen with N-ethyl-N-nitrosourea (ENU) (22). Mice were bred and housed at the Faculty of Medicine, Chulalongkorn University. All experiments were performed with the approval of the Animal Experimentation Ethics Committee of Chulalongkorn University Medical School with all relevant institutional guidelines**.**

**Pristane-induced lupus mouse model**

The C57BL/6 WT and *Sting*$^{gt/gt}$ mice (8-10 weeks old) were injected with the acyclic saturated hydrocarbon oil, tetramethylpentadecane (TMPD), or pristane (#P2870, SIGMA-ALDRICH Co., MO, USA). Two groups of mice were injected with 500 µl of 0.2 micron-filtered pristane via the intraperitoneal (IP) cavity (4). Blood samples were collected before pristane induction and every two months after the intraperitoneal injection. The mice were monitored for clinical symptoms and humanely sacrificed if they showed distressing symptoms.

**Sample collection and preparation**

The organs were harvested from the mice after six months of pristane injection. The splenocytes were isolated, and cold ACK lysis buffer was added to lyze red blood cells for 5 minutes. The splenocytes were washed and resuspended in 0.5% BSA in PBS for flow cytometry. Kidneys were fixed with 4% Paraformaldehyde in PBS. Lungs were perfused with normal buffer formalin to inflate the lungs and fixed with 4% Paraformaldehyde in PBS for tissue pathology.

**Detection of anti-dsDNA by enzyme-linked immunosorbent assay (ELISA)**

Using the previous protocol, the quantitative ELISA for anti-dsDNA was performed from the sera collected six months after pristane injection. In brief, 100 ul of 10 ug/ml dsDNA (UltraPure™ Calf Thymus DNA Solution, Invitrogen, USA) were incubated in a 96-well plate at 4˚C overnight. The plates were washed five times with 300 ul of 0.05% Tween in PBS (washing buffer) and blocked with 100 ul 0.1% Tween in 3%BSA/PBS (blocking buffer) at room temperature for 1.5 hours, then washed five times. The sera were diluted (1:100) in blocking buffer and incubated in the DNA-coated plate at room temperature for 1.5 hours. Sera were discarded and washed five times, then 100 ul of HRP-conjugated goat anti-mouse isotype-specific antibody (1:4000) was added, incubated at 37˚C for 1 hour, and then washed five times. The 100 ul of 1:1 ABTS peroxidase substrate solution A and B mixture were added at room temperature. ELISA reaction was stopped by adding 100 ul of 1N $H_2SO_4$. ELISA plate read the absorbance using a plate reader at wavelength 450 nm.

**Flow cytometry analysis**

Splenocytes were resuspended with staining buffer (0.5%BSA in PBS and 0.09% azide) to obtain the concentration of 20 x $10^6$ cells/ml. The splenocytes (1 x $10^6$ cells) were stained with flow antibody including CD4 (GK1. 5; #100423), CD8 (53-6. 7; #100708), CD62L (MEL-14; #104417), CD44 (IM7; #103035), CD3ε (145-2C11; #100312), ICOS (C398.4A; #313517), CD11c (N418; #117312), B220 (RA3-6B2; #103222), CD11b (M1/70; #101228), I-Ab (AF6-120.1; #116406), PDCA-1 (129c1; #127103), CD80

(16-10A1; #104733), GL7 (GL7; #144604), CD138 (281-2; #142506), F480 (BM8; #123112), Ly6c (HK1.4; #128022), CD45RB (C363-16A; #103307), Ly6c (HK1.4; #128022), Ly6g (1A8; #127608), IgM (RMM-1; #406512), IgD (11-26c.2a; #405718), CD21 (7E9; #123415), CD23 (B3B4; #101613), CD19 (6D5; #115522), IFNγ (XMG1.2; #505821), IL17A (TC11-18H10.1; #506921), and Gr1 (RB6-8C5; #108412), (Biolegend, San Diego, CA, USA), FAS (15A7; #12095181) (Invitrogen, Frederick, MD, USA). Cell viability (eBioscience™ Fixable Viability Dye eFluor™ 780, #65-0865-14, Thermo fisher scientific, CA, USA).

For cell surface marker staining, splenocytes were incubated with a staining master mix on ice or at 4˚C in the dark for 15 minutes. The stained cells were washed, resuspended with 200 ul of staining buffer, and centrifuged at 400 *rcf* for 5 min. The stained cells were fixed with 200 ul of fixing buffer (1%Paraformaldehyde in PBS). For intracellular staining, splenocytes were stained with surface marker antibodies and were fixed in 200 ul of fixation buffer (BioLegend, San Diego, CA, USA) at 4 °C overnight. Then, splenocytes were stimulated with PMA 25 ng/ml, ionomycin 1 ug/ml (Sigma-Aldrich, Darmstadt, Germany), and 1X GolgiPlug (brefeldin A, Biolegend, San Diego, CA, USA). The stimulated splenocytes were incubated in 5% $CO_2$, at 37˚C, for 4 hours. The surface-stained splenocytes were incubated with 1X permeabilization buffer (BioLegend, San Diego, CA, USA), resuspended in 50 ul of 1X permeabilization buffer, and performed intracellular cytokine staining. The flow cytometry was analyzed using BD™ LSR-II (BD Biosciences, USA) and FlowJo software (BD Biosciences, USA).

**Detection of the anti-nuclear antibody (ANA) by immunofluorescence assay**

Sera were diluted at 1:2000 in 1X PBS. Thirty microliters of diluted sera were applied to the slides coating with HEp-20-10 cells and primate liver (EUROIMMUN AG, Luebeck, Germany) and incubated at room temperature for 30 minutes. Then, slides were washed with 0.2% Tween-20 in PBS (washing buffer) and incubated with the fluorescein-labeled anti-mouse antibody (1:5000) for 30 minutes. The slides were washed with washing buffer for 5 minutes x 3 times and counterstained with diluted Evans Blue (10 drops: PBS 150 mL). The slides were embedded with a mounting medium and covered with the cover glass. The researcher will be blinded and grade the intensity as 4= maximal fluorescence (brilliant yellow-green), 3 = less brilliant (yellow-green fluorescence), 2= definite (dull yellow-green), and 1= very dim (subdued fluorescence) (19).

**Immunohistochemistry**

The mice were euthanized six months after pristane injection. The whole lung was infused with paraformaldehyde to dilate the alveoli through the trachea. The trachea was clamped using thread and then immersed in 4% paraformaldehyde/PBS. The harvested kidneys and lungs were fixed with 4% paraformaldehyde/PBS and embedded in paraffin. The 5 µm of tissue sections were stained with Hematoxylin and eosin (H&E). The clinicians blindly graded the H&E stained sections to score the histopathology of each organ. The kidney scores were graded as glomerular and interstitial scores previously described (30). The lung pathology was evaluated as diffuse pulmonary hemorrhage and interstitial inflammation (31, 32).

**Study population**

Blood samples from 173 healthy donors were collected from the blood bank at Ramathibodi hospital. The blood of 302 SLE patients who followed up at the Rheumatology clinic, Ramathibodi Hospital, from 2016 to 2021 were collected. All patients were older than 18 and met the 1997 ACR criteria (1) or SLICC criteria 2012 (2) for SLE classification. The exclusion criteria are SLE patients with a history of cancer. The medical records were reviewed and analyzed if the patients had followed up at Ramathibodi Hospital for at least five years since the diagnosis. The study (MURA2015/731and

MURA2021/177)) was approved by the Faculty of Medicine Ramathibodi Hospital ethics committee and conducted according to the guidelines of the Declaration of Helsinki.

**Study design**

A cross-sectional study was conducted at the Division of Allergy, Immunology, and Rheumatology, Department of Medicine, Faculty of Medicine Ramathibodi Hospital. The inheritance patterns of each variant were identified and categorized for subgroup analysis. The clinical data were compared between the different mutation patterns. The primary endpoint was STING variants that increase SLE risk and the clinical manifestations associated with STING mutation in SLE patients.

**Clinical assessment**

Clinical data at diagnosis and five years of follow-up were collected. Disease activity at diagnosis, $1^{st}$ and $5^{th}$ year of follow-up was assessed by systemic lupus erythematosus disease activity index 2000 (SLEDAI-2K) (3), and remission was defined as clinical SLEDAI = 0 and physician global assessment <0.5 by the 2021 DORIS definition of remission in SLE (4). Disease flare was defined as SLEDAI-2K increase ≥3 (3). End organ damage in $5^{th}$ year of follow-up as measured by Systemic Lupus International Collaborating Clinic/American College of Rheumatology Damage Index (5).

**Identification of STING genotype in SLE patients**

PBMC were isolated using Lymphoprep™ (STEMCELL Technologies Inc., Vancouver, BC, Canada) and followed the manufacturing protocol. AllPrep® DNA/RNA/Protein Mini Kit (QIAGEN GmbH, Germany) has been used to isolate DNA, RNA, and protein from the samples according to manufacturing protocol. Isolated DNA from the buffy coat and peripheral blood mononuclear cells (PBMC) of healthy donors and SLE patients were genotyped by Real-time PCR using TaqMan™ GTXpress™ Master Mix (Applied Biosystems, Thermo Fisher Scientific Inc., Waltham, MA USA). We designed TaqMan probes to detect *STING* variants at c.212G>A (R71H, rs11554776, Assay ID C_62979_10), c.689G>C (G230A, rs78233829, Assay ID C_104371077_10), c.695G>A (R232H, rs1131769, Assay ID C_26015196_10), c.878G>A (R293Q, rs7380824, Assay ID C_28947918_10), and mutation at c.852G>T were designed and customized by the author (R284S, Assay ID ANXGV44). The TaqMan genotyping assay was performed using Bio-Rad CFX96 Touch Real-Time PCR (Bio-Rad Laboratories, Inc. CA, USA). The qPCR reactions were detected under the PCR condition of 95°C, 3 minutes for reaction activation followed by 95°C, 15 seconds, and 60°C, 1 minute for 40 cycles. The signal was acquired at 60°C with two channels of fluorescence dyes, FAM and HEX.

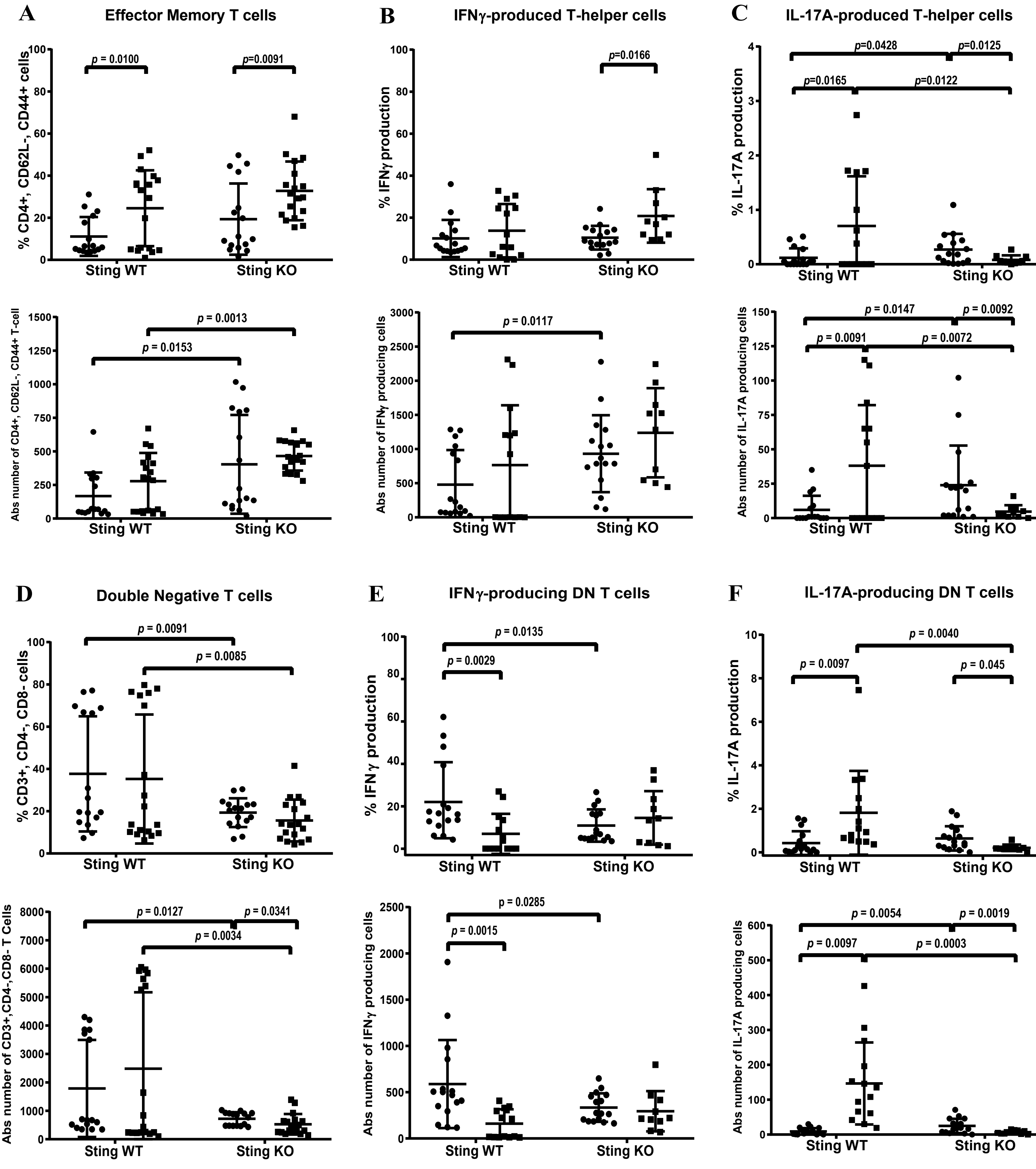


**Supplementary Figure 1. STING involved in double-negative T cell expansion and IL-17A production**
**The isolated splenocytes were stained and analyzed by flow cytometry. Data show the proportion and the absolute number of cells (A) effector memory T cells (CD4+, CD62L-, CD44+), (B) IFN-γ producing T helper cells (IFNγ+CD4+), (C) IL-17A producing T helper cells (IL-17A+CD4+), (D) double-negative T cells (CD3+CD4-CD8-), and (E) IFN-γ producing double-negative T cells (IFNγ+CD3+CD4-CD8-), and (F) IL-17A producing double-negative T cells. Data are shown as mean ± SEM; p-values indicated in the figures (N = 10-18 mice/group).**

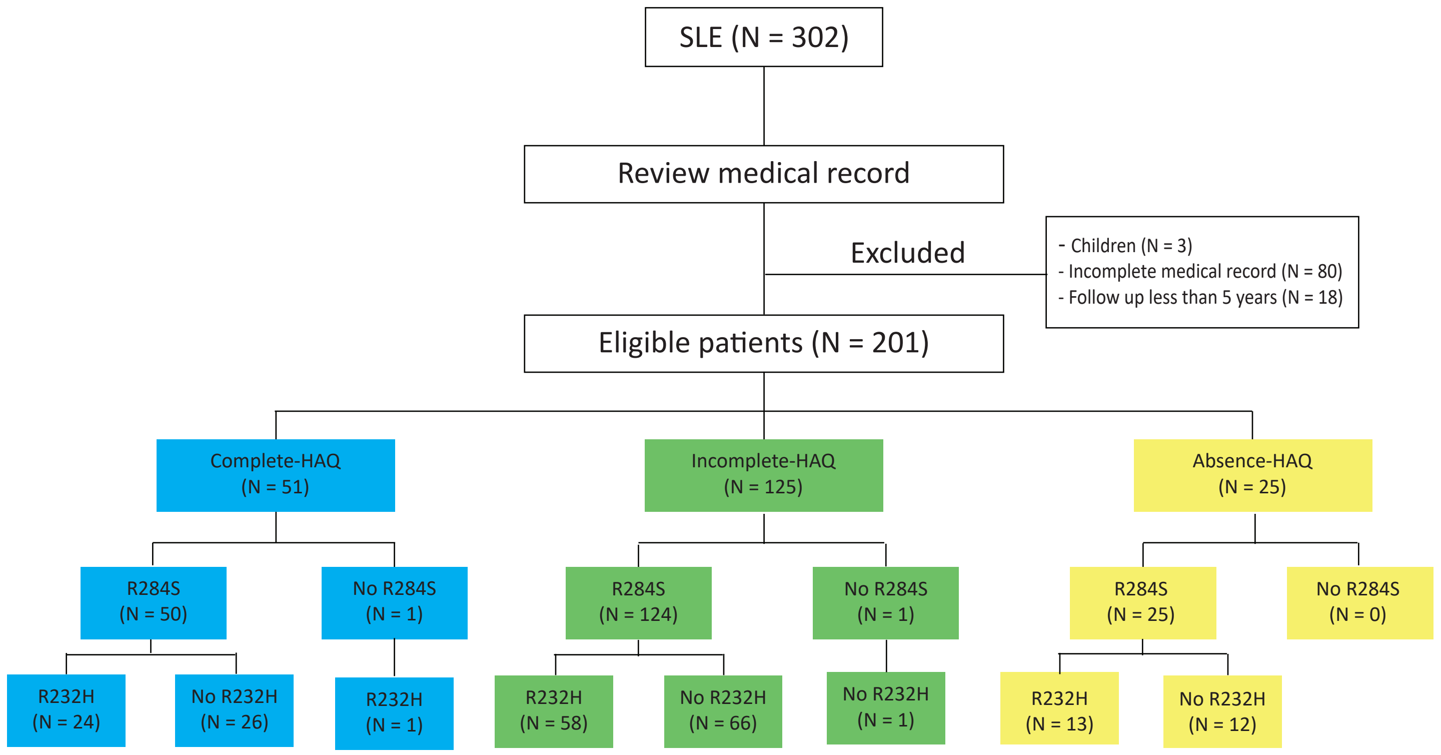


**Supplementary Figure 2.** Distribution of STING genotypes in clinical analysis of SLE patients
